# Clonal Hematopoiesis Increases the Risk of Autoimmune Hemolytic Anemia

**DOI:** 10.64898/2026.04.29.26351845

**Authors:** Stennio Da Silva Faria, Rafael Moisan, Estelle Lecluze, Thomas Pincez

**Affiliations:** CHU Sainte-Justine Azrieli Research Center, Montreal, Quebec, Canada; Faculty of Medicine, Université de Montréal, Montreal, Quebec, Canada; Division of Pediatric Hematology-Oncology, CHU Sainte-Justine, Montreal, Quebec, Canada

**Keywords:** Autoimmune hemolytic anemia, Clonal hematopoiesis, Immune thrombocytopenia

## Abstract

We show that clonal hematopoiesis is associated with an increased incidence of autoimmune hemolytic anemia. The hazard ratios of autoimmune hemolytic anemia and immune thrombocytopenia associated with clonal hematopoiesis were similar.

## Main

Autoimmune hemolytic anemia (AIHA) is a rare autoimmune disorder characterized by the immune-mediated destruction of red blood cells.^1^ AIHA can occur throughout life, but its incidence increases with age. AIHA can be secondary to lymphoid malignancies, systemic lupus erythematosus (SLE) or, more rarely, inherited error of immunity. In the absence of an underlying cause identified, AIHA is considered primary. Recently, clonal hematopoiesis (CH) has been identified as increasing the risk of immune thrombocytopenia (ITP),^2,3^ another immune cytopenia. CH is the age-related expansion of a somatic variant conferring a selective advantage to the hematopoietic stem or progenitor cell in which it occurs.^4^ Whether CH also increases the risk of AIHA is unknown. The existence of such risk and its magnitude cannot be extrapolated from ITP data for several reasons: 1) ITP has a higher incidence than AIHA,^1,5^ 2) AIHA and ITP pathophysiology have several differences,^1,5^ 3) the increase risk conferred by a disease can differ between AIHA and ITP (for example, some inherited error of immunity have been reported associated only with AIHA or with ITP).^6^

Here, we leveraged the large UK Biobank cohort (n=493,659) to assess the effect of CH on AIHA incidence. We used the published pipeline and results of CH curation available in the UK Biobank.^7^ This dataset includes high-confidence CH calling from the sample drawn at the inclusion in the UK Biobank, identifying 15,613 patients with CH (3.2%). From the 251 patients with AIHA, we included only those with primary incident AIHA. We excluded patients with an AIHA diagnosis made before inclusion and those with lymphoid malignancy or SLE diagnosed before AIHA to avoid confounding factors in CH occurrence due to underlying causes or treatment.

Of the 136 patients with primary incident AIHA, we identified 15 CH clones in 12 patients (8.8%). The CH-carrying genes were *DNMT3A* (n=7), *TET2* (n=3), *SRFS2* (n=2), *ASXL1* (n=1), *IDH2* (n=1), and *PPM1D* (n=1). The mean (± standard deviation) age at AIHA diagnosis was higher in patients with CH than in those without (63.8 ± 4.5 vs 59.7 ± 7.2 years, p=0.005). To analyze the association between CH and AIHA risk, we fitted a multivariate cox regression model adjusted on age, sex, and the first 10 principal components to consider genetic ancestry. We identified that CH was associated with a higher risk of AIHA (adjusted hazard ratio [aHR], 2.33; 95 confidence interval [CI] 1.28-4.23; p=0.006, Figure 1A). We then sought to investigate whether the magnitude of AIHA and ITP risk associated with CH were similar. We investigated the 572 patients with primary incident ITP (53 with CH [9.3%], Figure S1) using the same criteria as AIHA. We found that the risk of ITP is similar to that of AIHA (aHR, 2.60; 95 CI, 1.95-3.45; p=5.6×10^-11^). Patients with CH had a higher cumulative incidence of ITP than AIHA, in line with the higher incidence of this immune cytopenia in general population (Figure 1B). Finally, we found similar results in a sensitivity analysis using logistic regression in all patients with AIHA and ITP, including those with disease before inclusion and with prior lymphoid malignancy and SLE (i.e., prevalent cases, Figure 1C).

**Figure 1.**
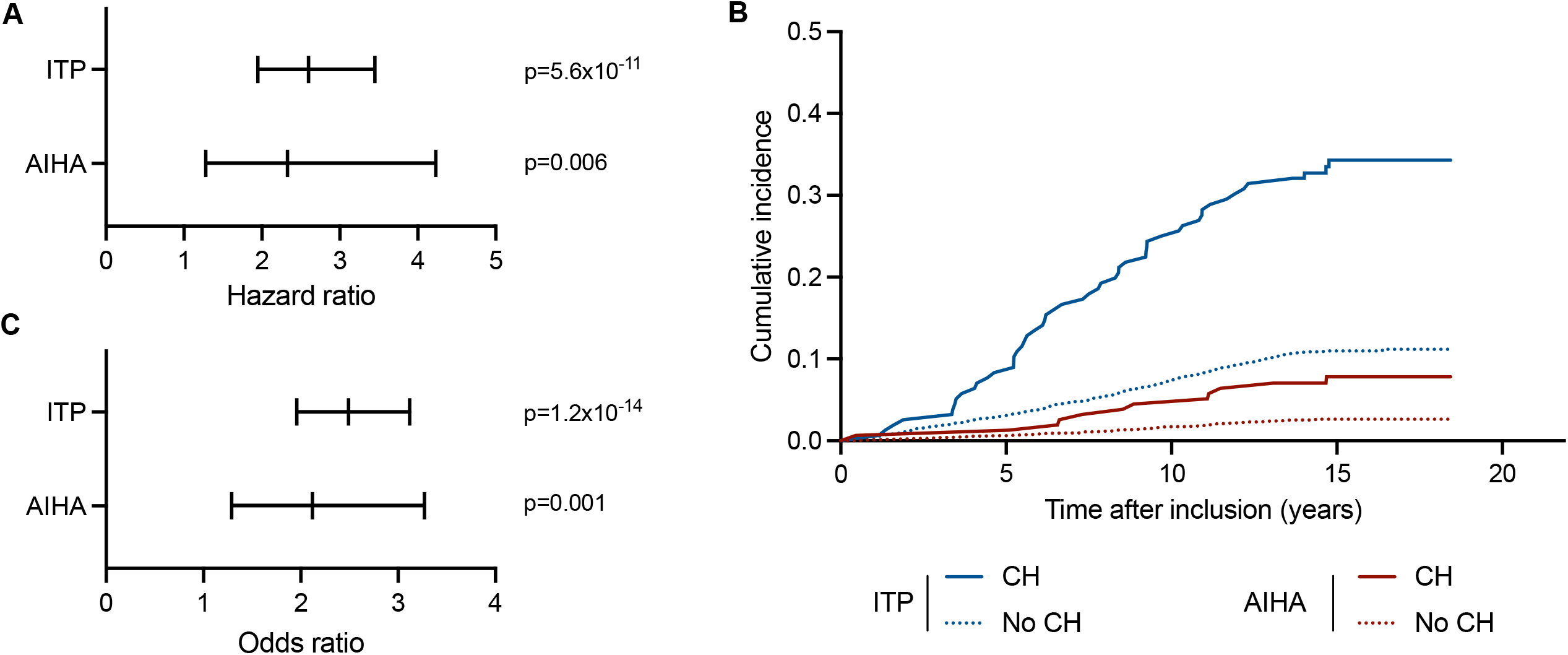
CH increases the risk of AIHA. A) aHR for the risk of incident primary AIHA (n=136) and ITP (n=572) from a multivariate Cox regression model adjusted on age, sex, and the first 10 principal components to consider genetic ancestry. B) Cumulative incidence of primary AIHA and ITP according to the presence of CH. C) Adjusted odds ratio for the risk of AIHA (n=251) and ITP (n=937) considering all cases (i.e., prevalent cases) from a multivariate logistic regression model adjusted on age, sex, and the first 10 principal components.

In summary, we found that CH was associated with a two-fold increased risk of AIHA. This risk conferred by CH appears to be of similar magnitude to the one of ITP. However, our sample size does not allow us to determine whether the slightly higher aHR for ITP is real or due to sampling fluctuations. We also found a similar proportion of patients with CH in both immune cytopenias. This proportion is lower than in one previous report of ITP, likely due to differences in population characteristics in terms of age and underlying disease.^2^ *DNMT3A* and *TET2* were the most frequent genes carrying CH in both AIHA and ITP, as in the general population.^3,4^ However, some differences may exist between the risk of ITP and AIHA associated with CH. In a previous study in the UK Biobank,^3^ the higher risk of ITP was conferred by *JAK2* variants, which were absent in our patients with AIHA. Our study has limitations. Although the UK Biobank is one of the largest existing biobanks,^8^ the rarity of AIHA limits the number of patients studied. This should be considered when interpreting our results and prevented us to study the gene-specific aHR. Finally, the mechanisms by which CH increases AIHA and ITP risk is unclear. Current knowledge suggest that autoimmunity is likely driven by inflammation and changes in immune function associated with CH.^9,10^

## Data Availability

All data produced in the present study are available upon application to the UK Biobank

## Conflicts of Interest Statement

T.P. has received research funding from Biossil Inc., unrelated to the topic. The other authors declare no competing financial interests.

## Acknowledgments

This research has been conducted using the UK Biobank Resource under Application Number 399545. This work uses data provided by patients and collected by the NHS as part of their care and support. The CHU Sainte-Justine Research Ethic Board approved the project.

## Notes

### Funding Statement

No funding received

### Author Declarations

The CHU Sainte-Justine Research Ethic Board gave ethical approval for this work (Project 2025-815)

## References

1. Michel M, Crickx E, Fattizzo B, Barcellini W. Autoimmune haemolytic anaemias. Nat Rev Dis Primers. 2024;10(1):82. doi:10.1038/s41572-024-00566-2

2. Fattizzo B, Marchetti A, Bosi A, et al. Clonal hematopoiesis in patients with autoimmune thrombocytopenia: an international multicenter study. Blood Advances. 2025;9(3):488–495. doi:10.1182/bloodadvances.2024014984

3. Liu Q, Wästerlid T, Smedby KE, et al. Clonal hematopoiesis of indeterminate potential and risk of immune thrombocytopenia. J Intern Med. 2025;297(6). doi:10.1111/joim.20092

4. Jaiswal S, Ebert BL. Clonal hematopoiesis in human aging and disease. Science. 2019;366(6465):eaan4673. doi:10.1126/science.aan4673

5. Hillier K, Kim T, Pincez T. Immunopathology of Immune Thrombocytopenia. Journal of Thrombosis and Haemostasis. Published online 9 February 2026. doi:10.1016/j.jtha.2026.01.011

6. Seidel MG, Hauck F. Multilayer concept of autoimmune mechanisms and manifestations in inborn errors of immunity: Relevance for precision therapy. Journal of Allergy and Clinical Immunology. 2024;153(3):615–628.e4. doi:10.1016/j.jaci.2023.12.022

7. Vlasschaert C, Mack T, Heimlich JB, et al. A practical approach to curate clonal hematopoiesis of indeterminate potential in human genetic data sets. Blood. 2023;141(18):2214–2223. doi:10.1182/blood.2022018825

8. Sudlow C, Gallacher J, Allen N, et al. UK biobank: an open access resource for identifying the causes of a wide range of complex diseases of middle and old age. PLoS Med. 2015;12(3):e1001779. doi:10.1371/journal.pmed.1001779

9. Kishtagari A, Corty RW, Visconte V. Clonal hematopoiesis and autoimmunity. Seminars in Hematology. 2024;61(1):3–8. doi:10.1053/j.seminhematol.2024.01.012

10. Wu H, Wei J, Yu Y, Wang N, Tan X. Clonal hematopoiesis of indeterminate potential and the risk of autoimmune diseases. J Intern Med. 2025;297(6):642–656. doi:10.1111/joim.20080

