## Supplementary material for "Clonal Hematopoiesis Increases the Risk of Autoimmune Hemolytic Anemia": Table S1

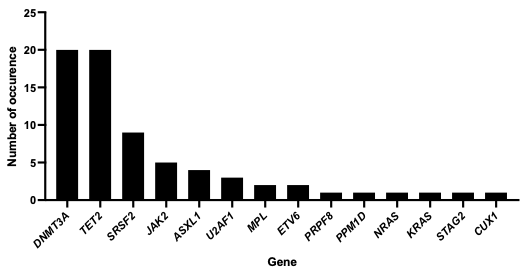

**Figure S1. CH-carrying genes identified in the 572 patients with incident primary ITP.** We identified 71 CH clones in 53 patients.
